# Comparison of actionable alterations in cancers with kinase fusion, mutation, and amplification

**DOI:** 10.1101/2024.05.24.24307868

**Authors:** Shinsuke Suzuki, Toshiaki Akahane, Akihide Tanimoto, Michiyo Higashi, Ikumi Kitazono, Mari Kirishima, Masakazu Nishigaki, Toshiro Ikeda, Shuichi Kanemitsu, Junichi Nakazawa, Erina Akahane, Hiroshi Nishihara, Kimiharu Uozumi, Makoto Yoshimitsu, Kenji Ishitsuka, Shin-ichi Ueno

## Abstract

Kinase-related gene fusion and point mutations play pivotal roles as drivers in cancer, necessitating optimized targeted therapy against these alterations. The efficacy of molecularly targeted therapeutics varies depending on the specific alteration, with great success reported for such therapeutics in the treatment of cancer with kinase fusion proteins. However, the involvement of actionable alterations in solid tumors, especially in relation to kinase fusions, remains incompletely understood. This study aimed to compare the number of actionable alterations in patients with tyrosine or serine/threonine kinase domain fusions, mutations, and amplifications. We analyzed 613 patients with 40 solid cancer types who visited our division between June 2020 and April 2024. To detect alterations involving multiple-fusion calling, we performed comprehensive genomic sequencing using FoundationOne^®^ companion diagnostic (F1CDx) and FoundationOne^®^ Liquid companion diagnostic (F1LCDx). Patient characteristics and genomic profiles were analyzed to assess the frequency and distribution of actionable alterations across different cancer types. Of the 613 patients, 44 had fusions involving kinases, transcriptional regulators, or tumor suppressors. F1CDx and F1LCDx detected 13 with kinase-domain fusions. We identified 117 patients with kinase-domain mutations and 58 with kinase-domain amplifications. The number of actionable alterations in patients with kinase-domain fusion, mutation, or amplification (median [interquartile range; IQR]) was 2 (1–3), 5 (3–7), and 6 (4–8), respectively. Patients with kinase fusion had significantly fewer actionable alterations than those with kinase-domain mutations and amplifications. However, those cancers with fusion involving tumor suppressors tended to have more actionable alterations (median [IQR]; 4 [2–9]). Cancers with kinase fusions tended to exhibit fewer actionable alterations than those with kinase mutations and amplifications. These findings underscore the importance of detecting kinase alterations and indicate the pivotal role of kinase fusions are strong drivers of cancer development, highlighting their potential as prime targets for molecular therapeutics.

## Introduction

The ability to determine complete exon sequencing of relevant tumor driver, suppressor, and resistance genes is paramount for optimizing personalized medicine. Comprehensive genome sequencing (CGS) can serve as a valuable detection tool for actionable alterations, which provide the biological basis of tumorigenesis. Kinase-related gene fusions, point mutations, and amplifications play an important role as drivers in cancer, necessitating optimized targeted therapy against these alterations. The FoundationOne^®^ companion diagnostic (F1CDx) and FoundationOne^®^ Liquid companion diagnostic (F1LCDx) are tissue- and blood-based broad companion diagnostics that have been approved by the Food and Drug Administration and clinically and analytically validated for all solid tumors. In Japan, F1CDx and F1LCDx are covered by public health insurance only after completion of standard treatment, which differs from practices in other countries [1].

Gene fusion, particularly those involving tyrosine kinase, plays a pivotal role as a driver mutation in the development of cancer. A hallmark example is the Philadelphia chromosome 9–22 translocation, characteristic of chronic myelogenous leukemia (CML), which generates the fusion protein BCR–ABL1 [2,3]. CML, driven by the tyrosine kinase gene fusion *BCR– ABL1*, stands out as the tumor with the most notable success in molecularly targeted therapeutics to date. Remarkably, almost 11 years of follow-up have shown that the efficacy of imatinib, a molecularly targeted drug, persists over time [4].

The prognosis of patients with non-small cell lung cancer (NSCLC) harboring oncogenic driver-gene alterations, such as epidermal growth factor receptor (EGFR) kinase mutation, anaplastic lymphoma kinase (ALK) fusion, c-ros oncogene 1 (ROS1) fusion, or rearranged during transfection (RET) fusion, and those with pan-cancer harboring neurotrophic tyrosine receptor kinase genes 1/2/3 (NTRK1/2/3) fusion, as well as those with cholangiocarcinoma harboring fibroblast growth factor receptor 2 (FGFR2) fusion has improved prominently with the advent of molecularly targeted drugs. However, the therapeutic effects of tyrosine kinase inhibitors (TKIs) vary between EGFR mutation and ALK, ROS1, RET, NTRK1/2/3 FGFR2 fusion. A tail-plateau, which is synonymous with durable response on a Kaplan–Meier survival curve, represents an attractive effect of molecularly targeted therapies [4] and immune checkpoint inhibitors [5,6] and has great clinical significance. In patients with advanced NSCLC with EFGR-mutations, not only the first generation of gefitinib but also the third generation of osimertinib, which is effective against the resistance mutation EGFR T790M, limited the durability of the response and did not show the benefit of tail-plateau on progression-free survival (PFS) curves [7,8]. However, TKIs in NSCLC cases with ALK [9] [10], ROS1 [11], RET fusion [12] and in pan-cancer with NTRK1/2/3 fusion [13] exhibited a tail plateau on the PFS curve. Further, Gao et al. analyzed a cohort of 9,624 samples from The Cancer Genome Atlas with 33 cancer types to detect gene fusion events. They focused on kinase fusion and showed that tumors with fusion events tend to have a lower mutational burden [14].

In this study, we hypothesized that the smaller the number of actionable alterations, the stronger the tumorigenesis and that the efficacy of targeted therapy would depend on the driver kinase alterations. Therefore, we aimed to compare the number of actionable alterations in patients with tyrosine or serine/threonine domain fusions, mutations, and amplifications using tissue- and blood-based CGS.

## Materials and Methods

### Patients

In total, 613 patients with 44 different solid cancer types visited our division between June 2020 and April 2024. Of these, 504 patients (82.2%) and 109 patients (17.8%) were analyzed using tissue- and blood-based CGS, respectively. All patients had solid cancer with advanced stage, with performance status of 0 or 1, and had completed standard treatment covered by Japanese health insurance. This cohort also included patients with untreated rare cancers and sarcomas lacking standard treatment protocols. We did not exclude any patients tested by these assays during the data collection period, except patients who were analyzed by F1LCDx and received anti–epidermal growth factor receptor (EGFR) antibody therapy. This exception was due to blood-based sequencing identifying multiple novel mutations, copy gains, and fusions associated with anti-EGFR therapy that frequently co-occur as subclonal alterations in the same patient [15]. We are reporting a retrospective study of medical records. The data were accessed for research purposes on 19 May 2024. We had access to information that could identify individual participants during or after data collection. These 40 cancer types were adenoid cystic carcinoma, bladder urothelial carcinoma, brain low grade glioma, breast invasive carcinoma, cervical squamous cell carcinoma and endocervical adenocarcinoma, cholangiocarcinoma, colon adenocarcinoma, esophageal carcinoma, glioblastoma multiforme, head and neck squamous cell carcinoma, kidney renal clear cell carcinoma, kidney renal papillary cell carcinoma, lung adenocarcinoma, lung squamous cell carcinoma, mesothelioma, neuroendocrine tumor, NUT carcinoma, ovarian serous cystadenocarcinoma, pancreatic adenocarcinoma, pheochromocytoma and paraganglioma, porocarcinoma, prostate adenocarcinoma, sarcoma, skin cutaneous melanoma, stomach adenocarcinoma, testicular germ cell tumor, thymoma, thyroid carcinoma, uterine carcinosarcoma, uterine corpus endometrial carcinoma, and uveal melanoma. Patient characteristics such as sex, age, smoking, heavy drinking (approximately 60g or more of pure alcohol per day average), prior chemotherapy, prior targeted therapy, and F1CDx or F1LCDx analyses have all been postulated to affect the number of actionable alterations (Table 1).

**Table 1.** Baseline characteristics of different subgroups (n=197)

|  | **Kinase alteration** |  |  | **Fusion**<br>**tumor**<br>**suppressor** | **p-value** |
| --- | --- | --- | --- | --- | --- |
| **Factor** | **Fusion** | **Mutation** | **Amplification** |  |  |
| n | 13 | 117 | 58 | 9 |  |
| Female sex, no.<br>(%) | 5 (38.5) | 64 (54.7) | 25 (43.1) | 2 (22.2) | 0.14 |
| Median age (SD) | 56.46<br>(19.21) | 61.22<br>(13.92) | 59.34 (13.11) | 67.78<br>(12.14) | 0.245 |
| Never smoked<br>(%) | 7 (53.8) | 62 (53.0) | 25 (43.1) | 4 (44.4) | 0.636 |
| Heavy drinking<br>(%) | 1 (7.7) | 17 (14.5) | 10 (17.2) | 2 (22.2) | 0.768 |
| Without prior<br>chemotherapy<br>(%) | 8 (61.5) | 75 (64.1) | 43 (74.1) | 7 (77.8) | 0.489 |
| Without prior<br>targeted therapy<br>(%) | 12 (92.3) | 104 (88.9) | 55 (94.8) | 9 (100.0) | 0.448 |
| blood-based CGS | 3 (23.1) | 13 (11.1) | 4 (6.9) | 2 (22.2) | 0.192 |
| (%) |  |  |  |  |  |
| Median no. (IQR)<br>of actionable<br>alteration | 2 (1- 3) | 5 (3-7) | 6 (4- 8) | 4 (2- 9) | <0.001 |

CGS, comprehensive genome sequencing; IQR, interquartile range.

### Ethics approval

This study was conducted in accordance with the Declaration of Helsinki and approved by the Ethics Committee and Institutional Review Board of Kagoshima University (approval number 180053). All patients provided written informed consent.

### Sequencing

Next-generation sequencing was performed using F1CDx and F1LCDx, which are CGS approaches involving the hybrid capture method. These tests targeted a panel of 324 genes, identifying base substitutions, insertions, deletion mutations, and copy number alterations in 309 genes; gene fusion in 36 genes; and tumor mutational burden (TMB) (a measure of the number of somatic protein-coding base substitutions and insertion/deletion mutations) in a tumor specimen. Protein tyrosine kinase and serine/threonine kinase were analyzed for kinase activating mutations. Amplification of CDK4/6, which coexists with other kinase alterations, was excluded from kinase amplification in this study. SS18-SSX and NAB2-STAT6 are disease-specific gene-fusion abnormalities and were detected separately via reverse-transcription polymerase chain reaction.

### Actionable alterations

All the detected gene alterations in cancer-related genes were annotated and curated using the COSMIC (https://cancer.sanger.ac.uk/cosmic), ClinVar (https://www.ncbi.nlm.nih.gov/clinvar/), CIViC (https://civicdb.org/home), SnpEff (https://www.accessdata.fda.gov/cdrh_docs/pdf17/P170019B.pdf.), and Clinical Knowledgebase (CKB) (https://ckb.jax.org/) databases. We calculated the validation (database; score) for mutation or fusion and the validation (score) for amplification or the clone status using PleSSision (Mitsubishi Space Software Co., Ltd., Tokyo, Japan), an outsourcing clinical sequencing system [10]. Mutations with a total score of 2 points or more were considered as actionable alterations. We used the following score tables:

1. for mutation or fusion of oncogenes: well-known driver status (more than 100 reports in COSMIC or pathogenic in ClinVar; 2), gain of function (CKB; 2), likely gain of function (CKB; 1.5), computational prediction of damage (SnpEff; 0.5);
2. for mutation or fusion of tumor suppressor gene (TSG): germline loss of function (gLOF) (pathogenic in ClinVar; 2), gLOF (truncate mutation; 2), gLOF (CKB or likely pathogenic in ClinVar; 2), gLOF of computational prediction (SnpEff; 0.5), somatic loss of function (sLOF) (pathogenic in ClinVar; 1), sLOF (truncate mutation; 1), sLOF (CKB or likely pathogenic in ClinVar; 1), computational prediction of damage (SnpEff; 0.5);
3. for amplification of an oncogene: neutral (0), copy number (CN)<4 (0), CN ≥4 (1), CN ≥8 (2),
4. for deletion of TSG, neutral (0), loss of heterozygosity (LOH) (1), uniparental disomy (UPD) (1), homologous deletion (HD) (2);
5. for clone status: main clone (1), subclone, tumor content >50% (1), subclone (0), uncertain (0), not a cancer clone or inconsistent with pathology (0), not inherited by cancer clones (-0.5).

For example, the TP53 R282W (LOH) mutation is annotated as sLOF (pathogenic in ClinVar; 1), LOH (1), and main clone (1), with a cumulative score of 3. This would be considered an actionable alteration, as our criteria specify that mutations with a cumulative score ≥ 2 are actionable alterations.

A multidisciplinary team comprising medical oncologists, pathologists, clinical laboratory technologists, bioinformaticians, and clinical geneticists conducted a comprehensive analysis to ascertain the clinical significance of these gene alterations.

### Statistical analysis

For number of actionable alterations, data are presented as median and interquartile range. Statistical significance was determined using analysis of covariance and the Steel– Dwass multiple comparison test. Results were considered significant at *P* < 0.05.

## Results

### Detection of fusion-genes in kinases, transcriptional regulators, and tumor suppressors

We analyzed 613 patients with 40 solid-cancer types via CGS. Fig 1 presents the distribution of patients with each cancer type. We identified 44 patients (9.6%) with fusions involving kinases, transcriptional regulators, or tumor suppressors. F1CDx and F1LCDx detected 13 patients (2.1%) with kinase-domain fusion (Table 2). In 12 of 13 patients with kinase-domain fusion, except for patient 9 with *CDK6* amp (CN=98), the other kinase-domain mutations and amplifications were mutually exclusive and did not coexist (Tables 1 and 3). Patient 13, with FGFR2-TACC2 fusion, exhibited coexistence with serine/threonine kinase mutation, *MAP3K13 p.I523V*(LOH), which was categorized as a variant of unknown significance. Additionally, patients 5, 6 and 11 showed coexistence of *MYC* amp (CN=9), *KRAS* p.*Q61H*, and *MYC* amp (CN=11), respectively, which is an interpreted oncogene (Table 3). Furthermore, all 13 patients demonstrated low TMB (Table 3).

**Fig 1.**
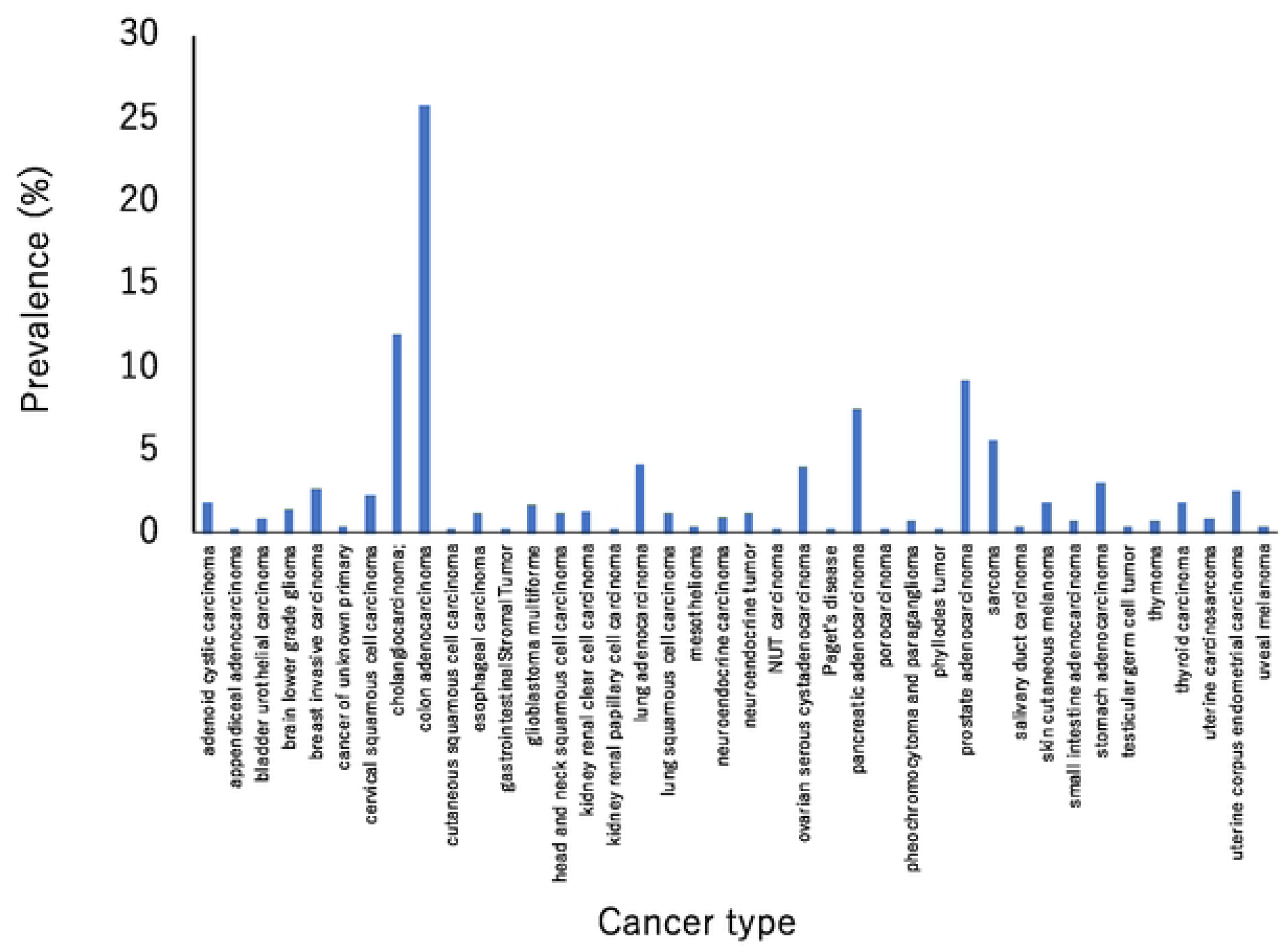
Proportions of cancer types in our sample. We analyzed the data of 613 patients with 40 different types of solid cancer using c tissue- and blood-based CGS.

**Table 2.**
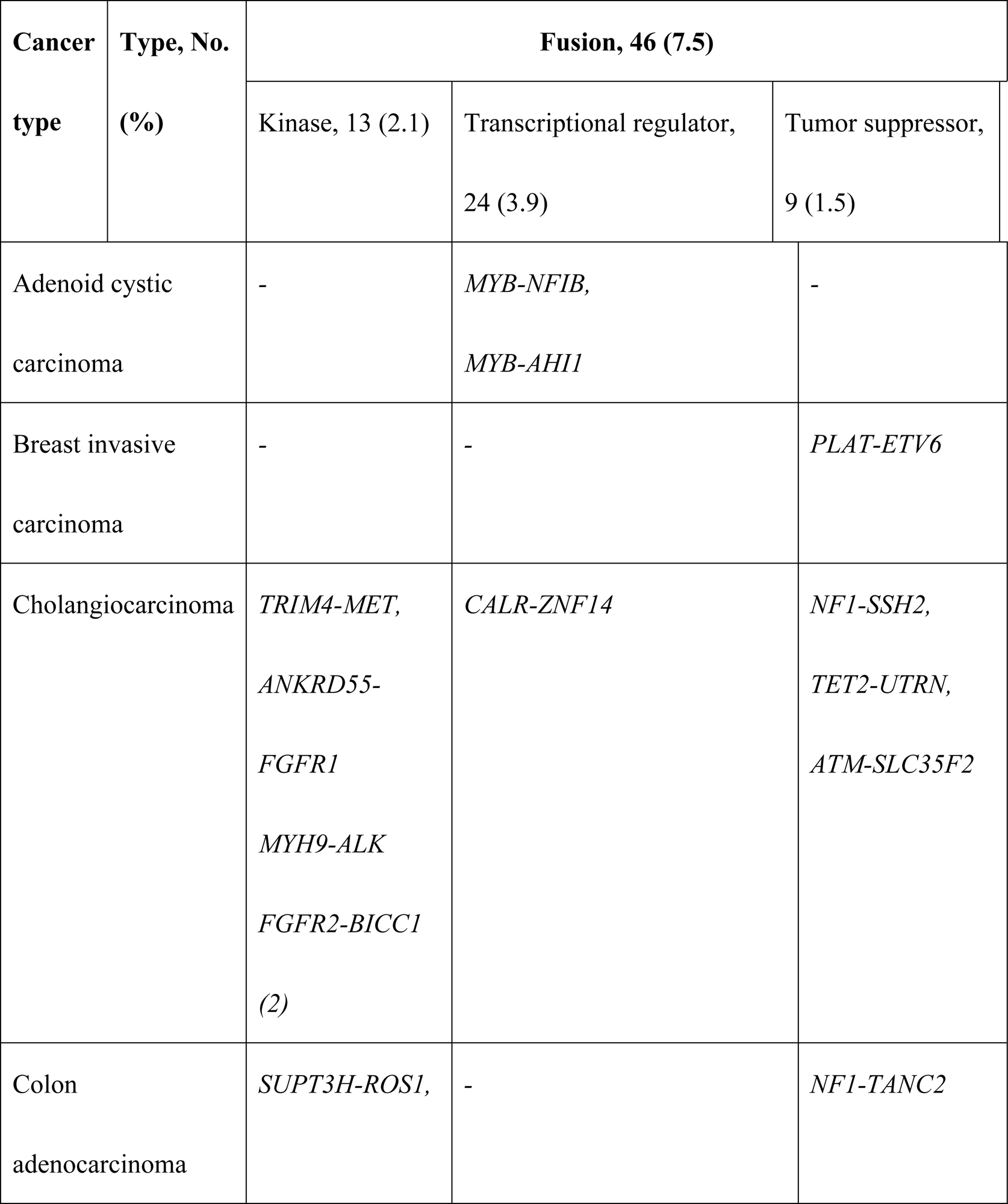
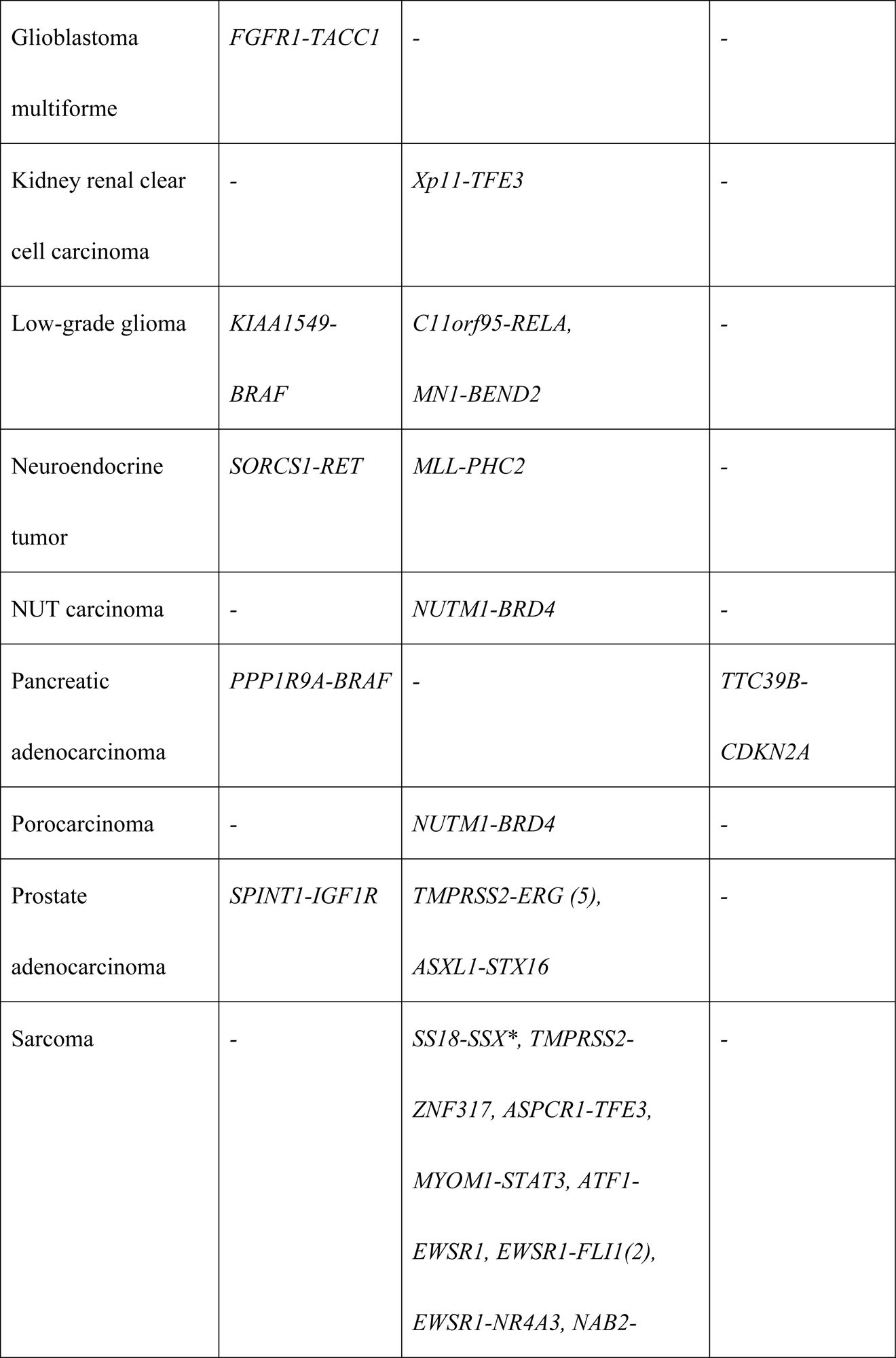
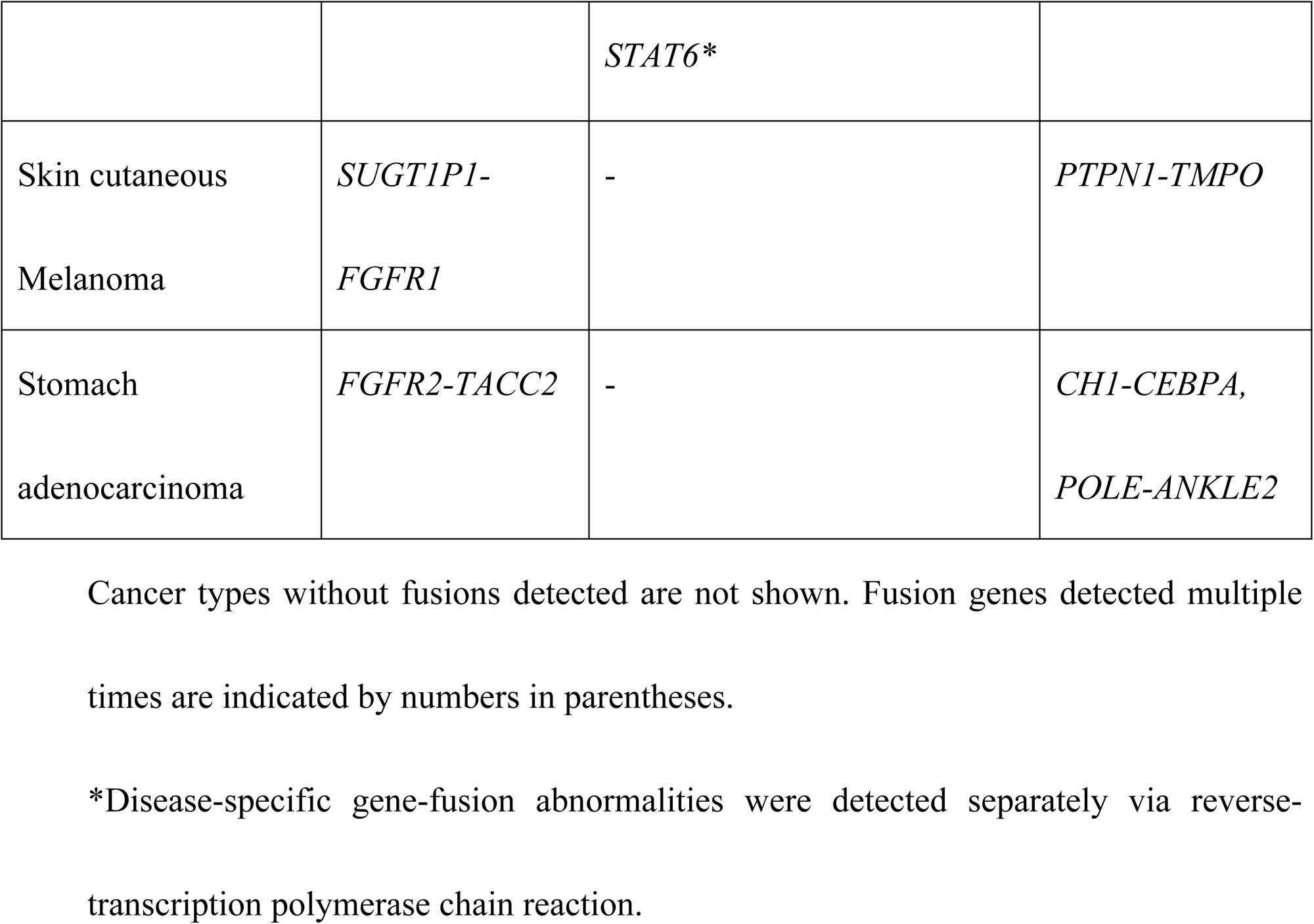
Fusion detection in cancer (n=613)

**Table 3.**
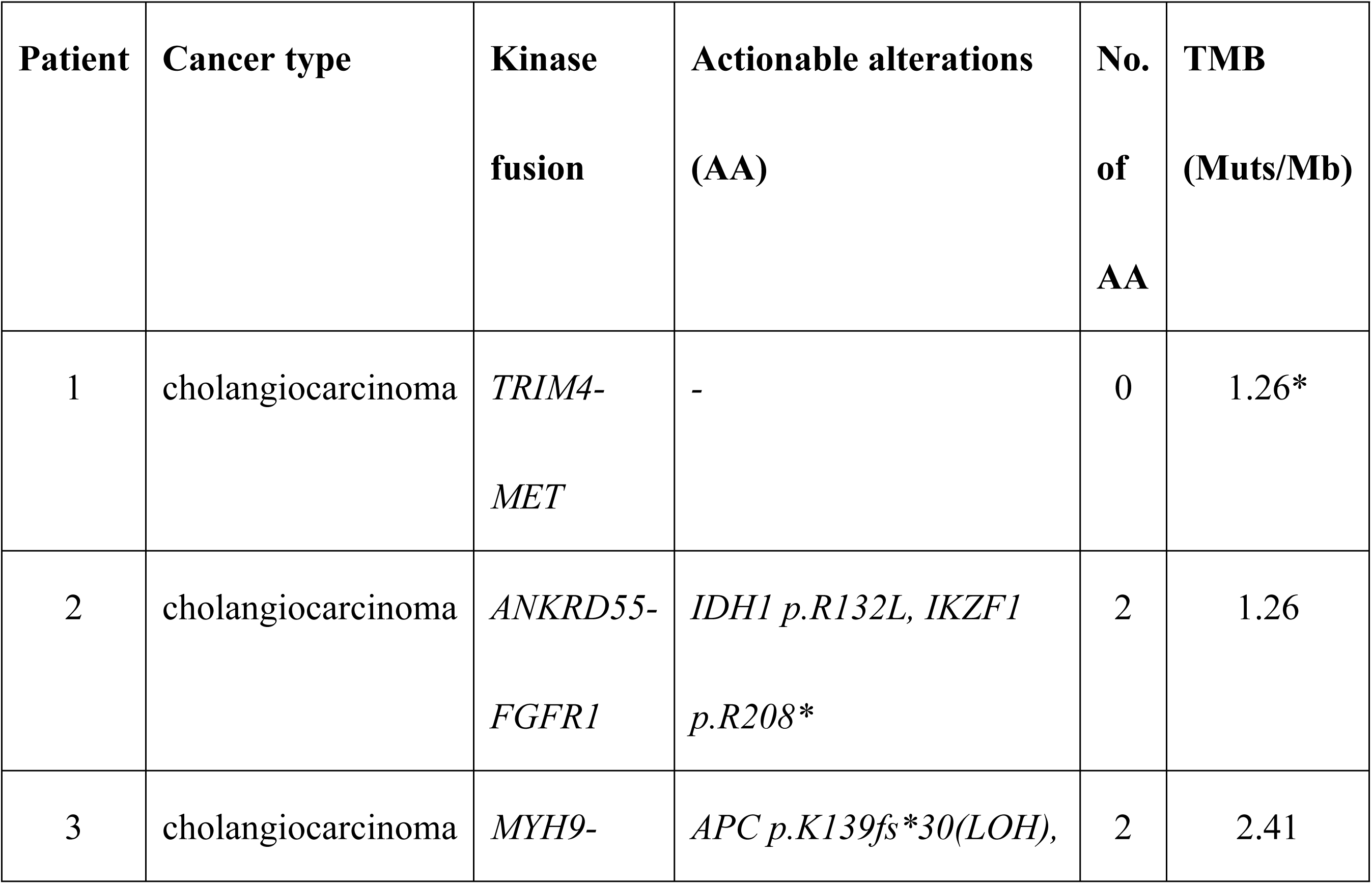
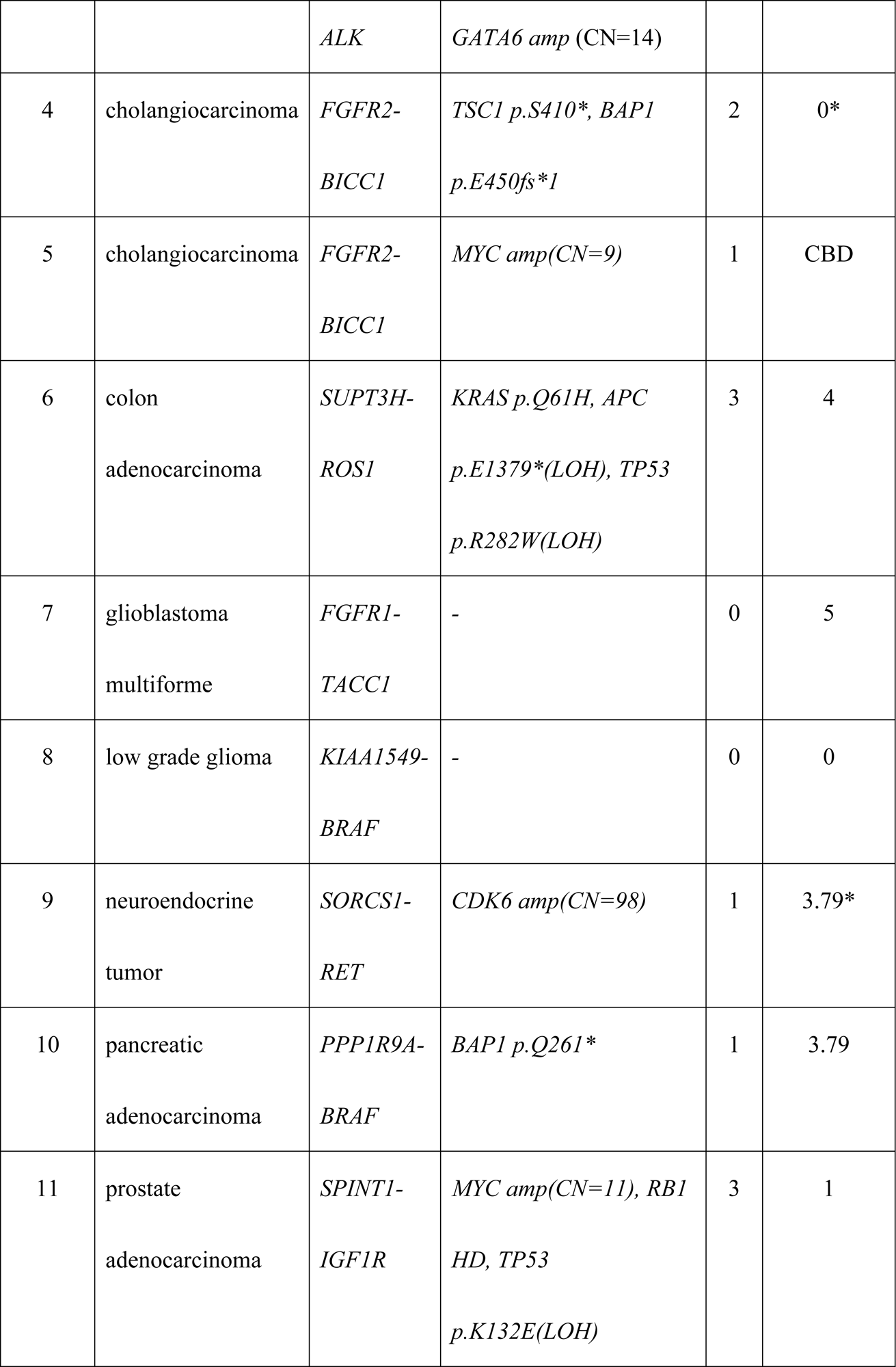
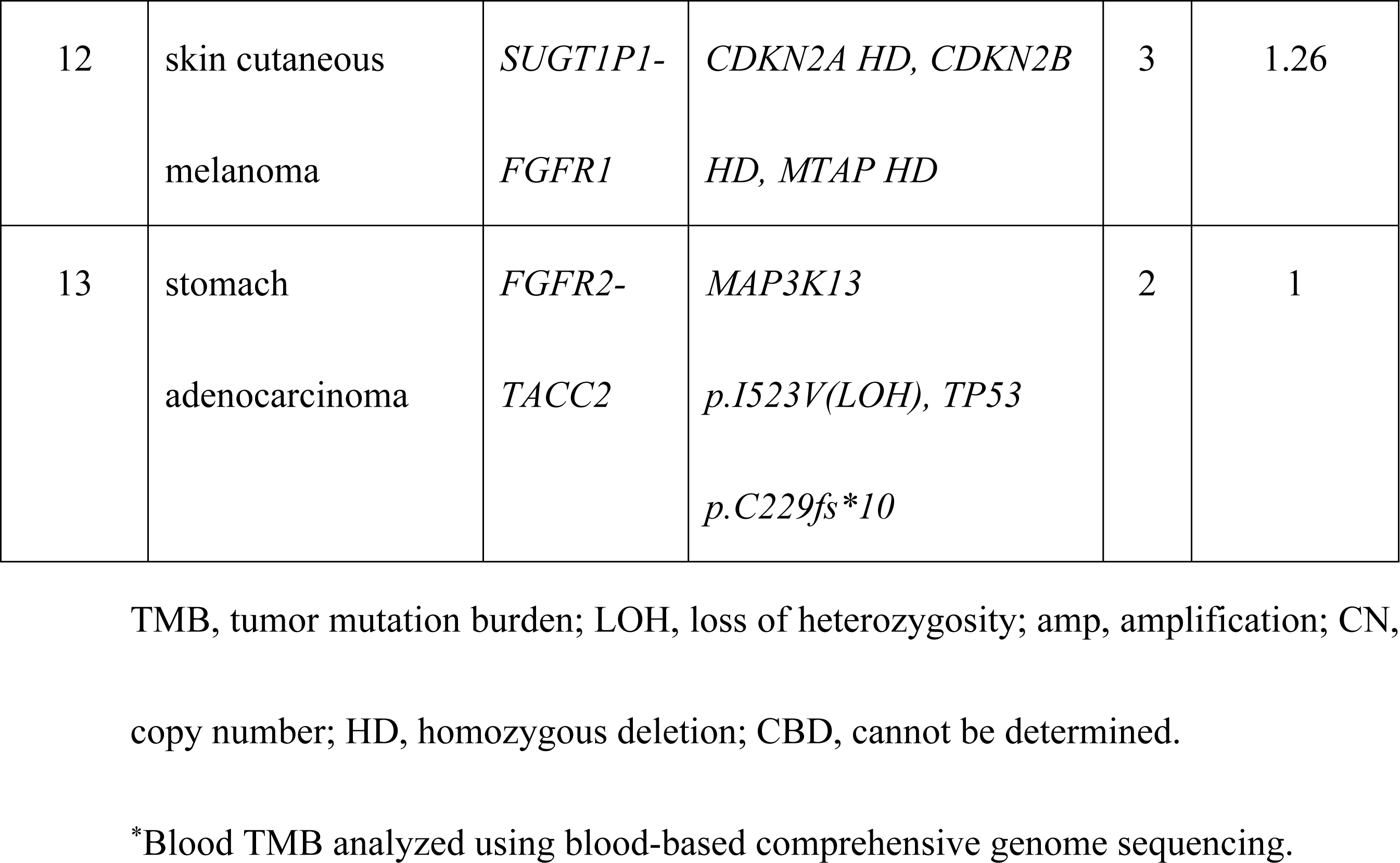
Actionable alterations of patients with kinase fusion (n=15)

We identified multiple complex chromosomal rearrangements involving *NUTM1*. These included a possible translocation between *NUTM1* on chromosome 15q and a region on chromosome 19p upstream of *BRD4* and a possible *BRD4-NUTM1* fusion in two patients (one with NUT carcinoma and one with porocarcinoma) (Table 2). Whether this fusion observed in porocarcinoma leads to the BRD4-NUTM1 fusion, which is characteristic of NUT carcinoma, remains unknown.

### Cancers with kinase fusion tended to have fewer actionable alterations

We identified 117 patients (19.1%) with mutations and 58 patients (9.5%) with amplifications involving the kinase domain (Table 1). After adjusting for baseline characteristics (sex, age, smoking, heavy drinking, prior chemotherapy and prior targeted therapy), we applied comprehensive genomic testing using F1CDx or F1LCDx to detect the frequency of actionable alterations in patients with kinase-domain fusions (n=13), mutations (117), amplifications (n=58), and suppressor fusion (n=9). The median counts (IQR) were 2 (1–3), 5 (3–7), 6 (4–8), and 4 (2–9), respectively (*P* < 0.001). The number of alterations was significantly lower in patients with kinase fusion than in those with kinase-domain mutations or amplifications (*P* <0.001; Steel–Dwass multiple comparison test) (Fig 2). However, the number of alterations was not statistically different for patients with suppressor fusions than for those with kinase-domain mutations and amplifications (Fig 2). In addition, the cancers with transcriptional-regulator fusion tended to have fewer actionable alterations **<u>2 (0–3.5)</u>** (median [IQR]).

**Fig 2.**
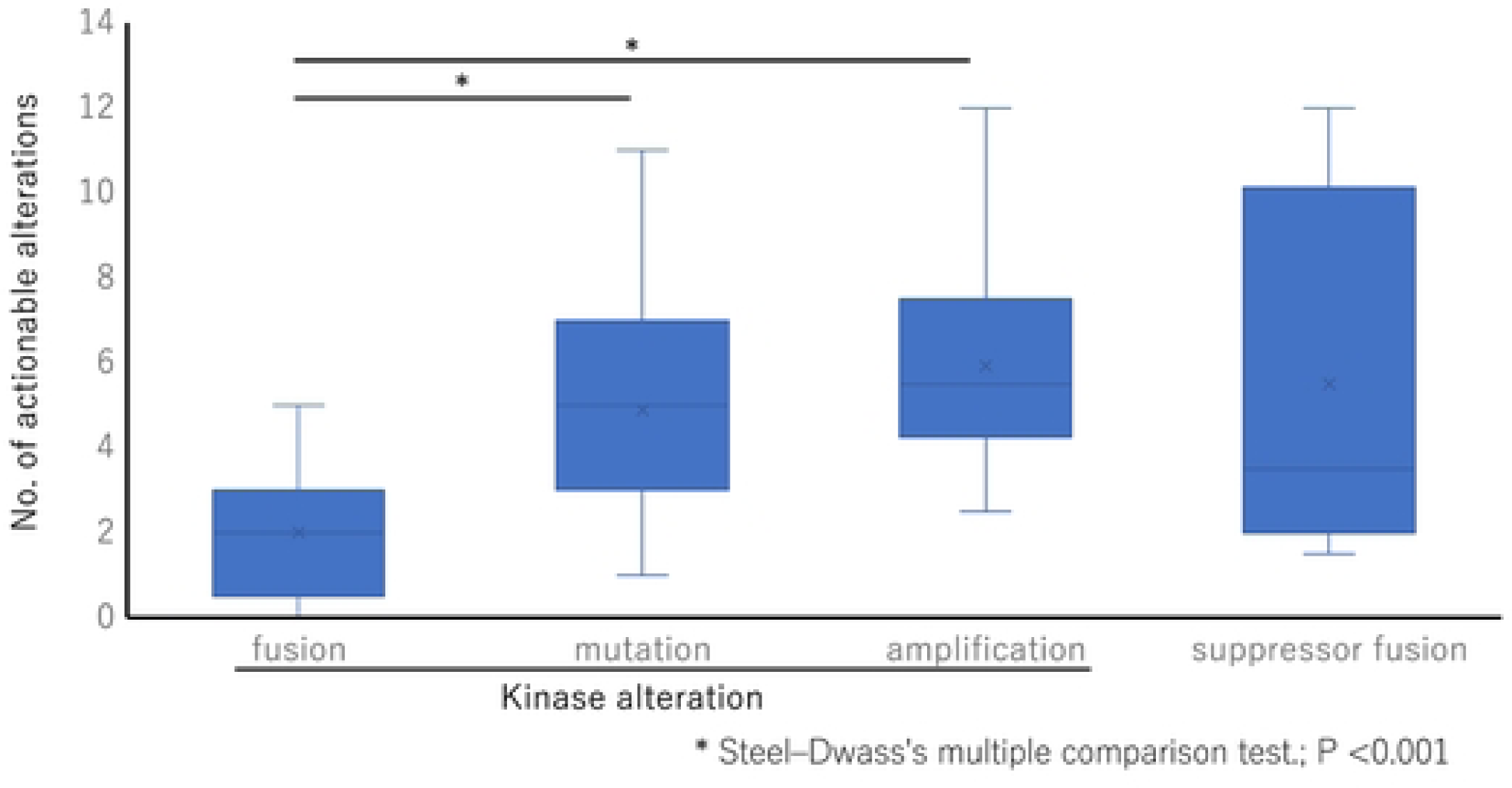
Distribution of actionable alterations across each alteration group. Patients with kinase fusions had significantly fewer actionable alterations than patients with kinase mutations and amplifications (*P* < 0.001; Steel–Dwass multiple comparison test). In contrast, fusions involving tumor suppressors other than kinase fusion did not show a significant difference in the number of actionable alterations.

## Discussion

These findings reveal that 7.5% of the 613 patients exhibiting 40 types of solid cancer patients included in the cohort had tumors with fusions. In particular, kinase fusions, which may have particular structural properties that are selected for during oncogenesis, were detected in 2.6% of the patients, accounting for 9 solid-cancer types. Cancers with kinase fusions tended to have fewer actionable alterations than those with kinase mutations and amplifications. Furthermore, the other kinase-domain mutations and amplifications were generally mutually exclusive in most patients with kinase fusions. These findings suggest that kinase fusion is a strong biological driver of cancer development. It is well established that CGS of tumor DNA is less sensitive than other methods for detecting fusions. This is a limitation of the study and requires further elaboration. Some tumors with fusions may have been missed, and data from those cases may have been included in other cohorts (for example, cases of kinase mutation only). Detection of fusion in DNA-sequencing data is difficult, and bimodal DNA- and RNA-based gene panels can be useful for this detection.

The presence of *IGF1R-SPINT1* in prostate adenocarcinoma, which we detected in patient 8, has not been previously reported. This fusion protein lacks an extracellular domain but retains the kinase domain. Removing the entire extracellular domain of the insulin-like growth factor (IGF) receptor activates the receptor, even without bound IGF [11]. Together with previously reported findings, this result suggests that the *IGF1R-SPINT1* fusion represents an activating mutation.

In patients 5, 6, 9, 11 and 13, we detected the coexistence of oncogene alterations with kinase fusions, which are considered generally mutually exclusive (Table 3). The amplification of *MYC* in patients 5 and 11 was limited to a small number of copies; CN=9 and CN=11, respectively. Furthermore, since the TSG (such as *RAD21* or *NBN*) in chromosome 8q, where MYC is present, was amplified with a same copy number, it was considered to be the result of polysomy of chromosome 8q. *KRAS p.Q61H* in patient 6 was interpreted as a conflicting interpretation of pathogenicity in ClinVar. *MAP3K13* (encoding LZK)-amplified head and neck squamous cell carcinoma cells harboring 3q gain are dependent on LZK expression for cell viability and colony formation [16]. LZK expression has been shown to be required for cell proliferation and anchorage-independent growth in MYC-overexpressing breast and hepatocarcinoma cell lines [17]. However, alterations that are predicted to inactivate LZK have also been reported in breast cancer [18]. Therefore, it is unclear whether *MAP3K13 p.I523V*(LOH) in patient 13 functions as a tumor suppressor or an oncogene. In these patients, the kinase fusions appear to be important for tumorigenesis even with the coexistence of the oncogene alterations. Although there is only one co-existing actionable alteration, high amplification of *CDK6 (CN=98)* was detected in patient 9. Sitthideatphaiboon et al. defined the pathways limiting EGFR-inhibitor response, including cell-cycle-gene, *CDK4/6* amplification [19]. In such patients, the effect of TKIs is expected to be influenced not only by the number but also by the type of comorbid actionable alterations.

Molecularly targeted drugs have been most successful in treating CML, which is driven by a kinase fusion (*BCR–ABL1*) [4]. Integrative genomic analysis revealed cancer-associated mutations in only three out of 19 patients (16%) who responded optimally to imatinib, whereas cancer-gene variants were detected in 15 of 27 patients (56%) with poor outcomes [20]. In our study, sole kinase fusion, without other actionable alterations, may be involved as a significant driver of the development of cancer in three patients (with cholangiocarcinoma, glioblastoma multiforme, and brain low grade glioma). However, in three patients with lung adenocarcinoma harboring an activating EGFR kinase mutation (EGFR p.E746_A750del, p.S768I, and p.L858R) without prior EGFR-targeted therapies, we detected 10, 4 and 3 actionable alterations, respectively. Moreover, all three patients had *TP53* loss-of-function mutations in uniparental disomy (data not shown). *TP53* mutations associate with faster resistance evolution in EGFR-mutant NSCLC and mediate acquisition of resistance mutations to EGFR TKIs [21]. Therefore, it is important to validate tumor alterations, including both driver and actionable mutations by CGS, to evaluate the efficacy of molecularly targeted therapy.

Focusing on kinase fusion, Gao et al. [14] showed that a significant proportion of patients harboring fusions involving cancer-driver genes had no driver-gene mutations. Moreover, their analysis highlights an important consideration for immunotherapy in patients with fusions. Specifically, the significantly lower mutational burden observed in patients with driver-gene fusions points toward a reduced efficacy of immunotherapy in these patients, despite fusion peptides being potentially good immunogenic targets. The TMB was also low in all the 13 patients with kinase-domain fusion. Based on their findings, Gao et al. [14] suggested that research into driver-gene fusion can result in the development of targeted drugs and immunotherapy.

This study has a limitation in terms of the small sample size. We only included 13 patients with kinase-fusion in the analysis; therefore, it is unlikely that the analysis was sufficiently powered to derive conclusions. Therefore, analysis of a large number of patients may be necessary.

## Conclusions

This study demonstrated that cancers with kinase fusion tend to have fewer actionable alterations than those with kinase mutations and amplifications, reflecting the strong dependence of tumorigenesis on kinase fusion. Therefore, detecting kinase mutations is crucial in developing molecularly targeted therapeutics and immune therapy.

## Data Availability

The data that support the findings of this study are available on request from the corresponding author, S.S. The data are not publicly available as the study participants did not consent to public sharing of their data.

## Authors’ contributions

SS designed and performed the experiments, analyzed the data, and wrote and revised the manuscript. TA and EA conducted the experiments. AT, MH, IK, MK, MN, TI, SK, JN, HN, KU, MY, KI, and SU analyzed the data and wrote and revised the manuscript.

